# Understanding and Mitigating Health Disparities Arising from the Social Determinants of Health

**DOI:** 10.1101/2023.08.23.23294496

**Authors:** Aaron Cho

## Abstract

Over the pandemic, health has become increasingly important, and our idea of what it means to be healthy has changed. Previously we viewed health disparities as an issue that affected those who were less fortunate, but the pandemic has shown us that the health of all of us is interconnected. Reducing health disparities is important for everyone, as it would improve health not just for those who are directly impacted, but for society as a whole. This study contributes towards reducing health disparities by analyzing what factors have the largest impact on health disparities, and discussing policy changes that are relevant to these factors. This study uses a random forest algorithm to identify important predictors of health from a large variety of factors in Chicagoland and the Bay Area. The analysis finds that race is an important factor in both Chicagoland and the Bay Area, and that lack of internet access and computing devices is an especially important predictor of neighborhoods with poor health.

## Introduction

The pandemic has increased everyone’s general health concerns. It has also changed our perspective about health. Prior to the pandemic, eating well and getting exercise were likely priorities for many people. However, as the pandemic unfolded, it became clear that one’s health is not simply a function of what a person does, but that our health is connected to the health of others. When others caught COVID-19, that mattered to their health as well as everyone they interact with. When one person gets the virus, it is bad for everyone, not just the person who gets it. This has become more and more apparent as the virus keeps mutating, causing new and more problematic variants to keep coming out. The pandemic has really reinforced that when it comes to health, everyone is in this together, and when one person is unhealthy, it is everyone’s concern. This is why it is an issue that it is much easier for some to maintain their health than others who may not be as fortunate. If we all want to stay healthy, we need to be concerned about how to keep us *all* healthy. That seems easy to understand, but one of the big stories out of the pandemic has been about health disparities.

Health disparities refer to the preventable difference of opportunity to be healthy between different segments of the population, and different types of people. During the pandemic, it was found that there was a racial health disparity. Lopez et al. (2021) conducted a study which discovered that Black, Hispanic, and Asian people have a much higher rate of infection from the virus, along with higher rates of hospitalization and death. Health disparities are not only found in race however, and can commonly be related to the environment one grows up in. Ray and Linden (2018) found that countries with bad health are commonly correlated with having low GDP per capita, and high GINI coefficients.

These factors that cause some places to be healthier than others are called social determinants of health. This brings up the question of what social factors contribute to making some neighborhoods more healthy than others, and what can be done to improve the areas which are less fortunate. This is especially interesting because these are areas where the government can take action. Once we have an idea on how to improve an environment, we can enact policy to change that environment. If for example, we found that high pollution levels, or low access to grocery stores were correlated with significantly less healthy neighborhoods, we would know to focus on reducing pollution or locating grocery stores in a more geographically equitable manner.

Since different environments create different health conditions, that implies that local conditions are important. Every place has different issues, and there is not one solution to fix the entire country. What works for one area of the country may be quite different than what would work for another area of the country. Drummer (2008) found that health was intrinsically linked with geography, and that spatial location plays a large role in shaping the environmental health factors and health risks. This study seeks to expand on all of these studies by identifying what specific factors are the most important to determining the health of a neighborhood in different areas. The focus is on the social determinants of health in two different geographical regions, the San Francisco Bay Area and Chicagoland. A random forest supervised machine learning algorithm is used to parse through a large selection of potential factors to identify the most important predictors of health in these two different areas and suggest ways in which policy changes can improve the health of not only these areas, but can have a cascading effect on the health of all of us.

### Literature Review

Others have studied the impacts of different neighborhood factors before. Many correlates have been found between health and its social determinants, as well as other similar factors. Education has been shown by past studies to be an important factor of health. For example, Braveman and Gottlieb (2014) found that life expectancy is strongly correlated with one’s level of educational attainment, and infant mortality rates are similarly correlated with the mother’s level of educational attainment. They also found that family income is correlated with lower rates of activity-limiting chronic disease, showing that poorer families tend to be less healthy. Braveman et al. (2010) found that in addition to poorer families having worse health, Blacks and Latinos tend to have lower educational attainment, and worse health as a result. Acevedo-Garcia et al. (2014) similarly found that the inequality between neighborhoods tends to have the largest effect on Blacks and Latinos. They found that in metropolitan areas, 40 percent of Black and 32 percent of Hispanic children live in very low opportunity neighborhoods, as compared to only 9 percent of White children.

In addition to education, it has also been shown that health is heavily impacted by the neighborhood one grows up in. Many studies show that neighborhood income plays an important role in how healthy we are. Kaplan et al. (2008) studied the correlation between income and psychological health, and found that all five scales of psychological health are strongly correlated with income. Marmot and Bell (2012) found that those who live in wealthier areas tend to have a longer life expectancy. They determined that in England, the difference in male life expectancy between the poorest and most affluent areas was above 9 years on average. Across countries however, the difference in life expectancy can be much larger. Marmot et al. (2008) found that the difference in average life expectancies for a girl born today can be more than 45 years, depending on the country she is born in.

In addition to affecting health, neighborhoods have also been shown to have a significant impact on upward mobility. Chetty et al. (2018) found using Census tract data, that “neighborhoods have substantial casual effects on children’s long-term outcomes at a granular level.” They examined how neighborhood measures of parent income, race, and gender, affected poverty and incarceration rates. In another study, Chetty et al. (2018) similarly found that neighborhoods matter using de-identified tax records. The study looked at two cities which had a large average income difference between children who grew up from birth there. The study then showed that kids who moved from the poorer city to the richer city on average improved their long-term income, increasing for each additional year they moved before age 23. Neighborhoods have many specific characteristics, and different studies think different characteristics matter more. Chetty et al. (2018) discovered that neighborhoods with high upward mobility tend to have the following five characteristics: less residential segregation, less income inequality, better primary schools, greater social capital, and greater family stability. In a separate study, Chetty et al. (2014) explored the quality of school can matter a lot, as replacing a teacher whose VA^1^ is in the bottom 5% with an average teacher would increase the present value of students’ lifetime income by approximately $250,000 per classroom.

Overall, past studies have identified many factors which are correlated with health, such as education level, race, and living environment, i.e., the neighborhood one grows up in. Many of these studies look primarily at the variables that are related to the wealth of neighborhoods. However, there are more things to study, such as other aspects of neighborhoods that impact health, and what factors play the largest role in impacting neighborhood health. This study takes inspiration from these studies, but since so much of health is local, I focus on 2 specific geographical regions. I compare and contrast the results from these different areas, and examine some general factors as well as specific factors that are most easily changed. This study is different from past studies in its focus and analysis. It adds to the literature by finding what aspects of neighborhoods we should focus on changing.

### Data

The data include a wide range of factors including race, income, and education level, along with access to different resources, such as vehicles, internet, stores and restaurants, and food assistance programs. All of the data are collected at the census tract level because census tracts are the lowest level at which the data are available for many of these factors. The dependent variable includes data on 11 different aspects of health and combines them to represent a general level of health. These data are from Centers for Disease Control and Prevention’s PLACES: Local Data for Better Health. My dependent variable represents a measure of health for a given census tract. This is created from the combination of the following 11 factors: 1) Percentage of arthritis among adults, 2) percentage of current asthma prevalence among adults, 3) percentage of high blood pressure among adults, 4) percentage of cancer among adults, 5) percentage of diagnosed diabetes among adults, 6) percentage of high cholesterol among adults who have been screened in the past 5 years, 7) percentage of chronic kidney disease among adults, 8) percentage of stroke among adults, 9) percentage of obesity among adults, 10) percentage of chronic obstructive pulmonary disease among adults, and 11) percentage of coronary heart disease among adults. Each census tract is given a health value which is calculated as the sum of these percentages. This means that census tracts with higher health values correspond to having worse health. The range of this variable is 40-233 with average values of 151 and 138 in Chicagoland and the Bay Area respectively. Figure 1 shows how this variable maps in Chicagoland (on the left) and the Bay Area (on the right).

**Figure 1.**
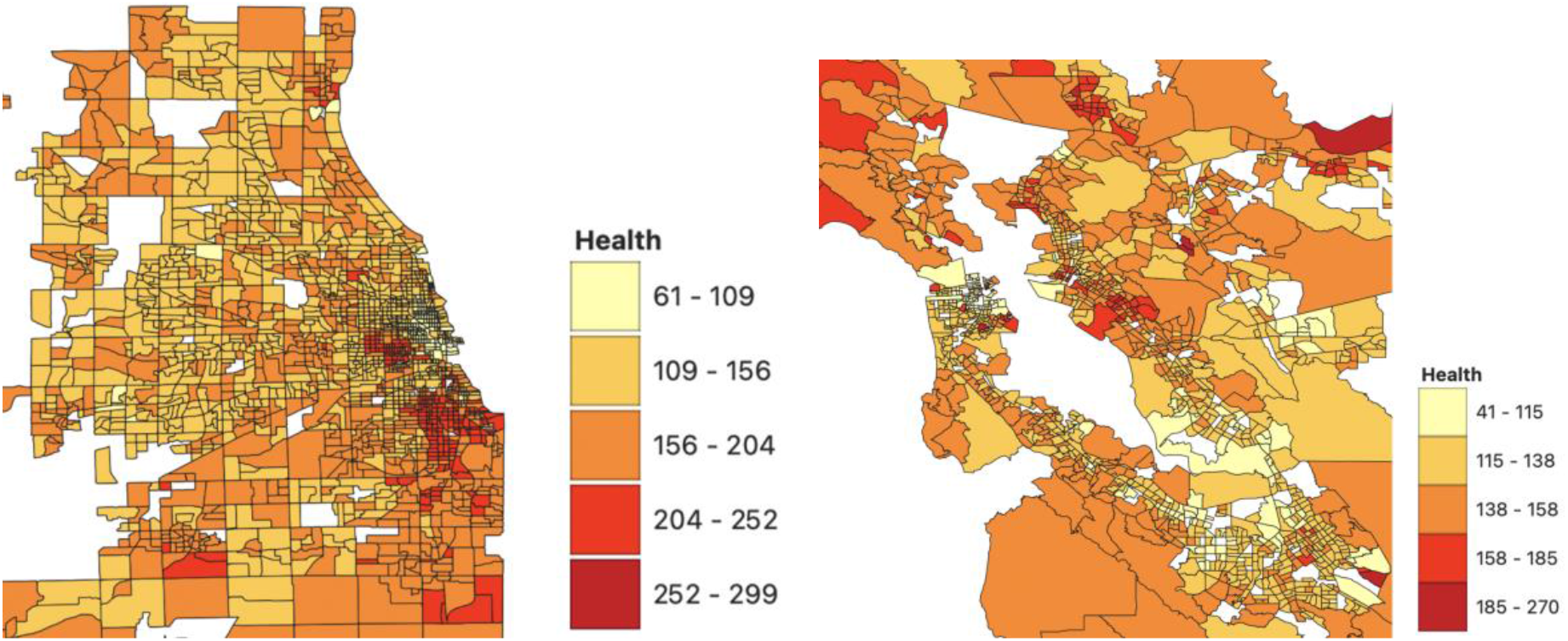
Health in Chicagoland (left) and the Bay Area (right)

As shown in the map, some areas are much healthier than others, raising the question of why this would be and what factors it may be related to. One might wonder, for instance, if the health patterns are related to race. Figure 2 shows how the Black population is dispersed in Chicagoland. While the two maps differ, some similarities are visually evident.

**Figure 2.**
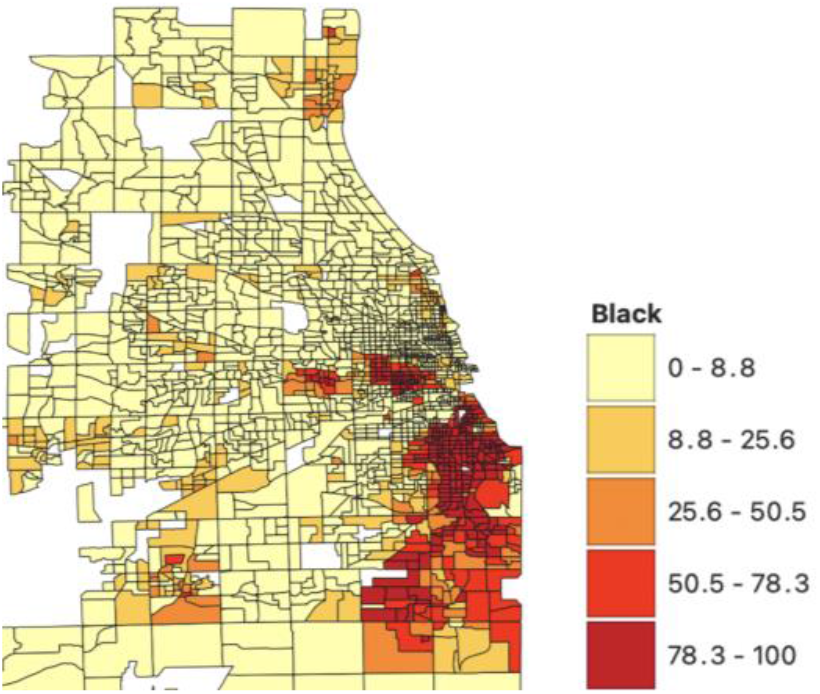
Percent Black in Chicagoland.

Of course, there are many possible explanations, so I explore the relationship with a large set of possible variables. I obtain my independent variables from a variety of sources, including the Department of Health and Human Services’ Agency for Healthcare Research and Quality’s Social Determinants of Health Database, the USDA Economic Research Service’s Food Environment Atlas, the CDC/ATSDR’s Social Vulnerability Index, the California Department of Public Health’s Modified Retail Food Environment Index, and the CDC’s National Environment Public Health Tracking Network. The variable names, their description, and the source are shown in Table 1.

**Table 1.** Independent Variables.

| Variable Name | Variable Description | Source |
| --- | --- | --- |
| ACS_TOT_POP_US_ABOVE1 | Total Population | ACS |
| ACS_PCT_ASIAN | Percent Asian | ACS |
| ACS_PCT_BLACK | Percent Black | ACS |
| ACS_PCT_HISPANIC | Percent Hispanic | ACS |
| ACS_PCT_WHITE | Percent White | ACS |
| ACS_PCT_HH_NO_COMP_DEV | No Computing Device | ACS |
| ACS_PCT_HH_NO_INTERNET | No Internet | ACS |
| ACS_GINI_INDEX | Gini Index | ACS |
| ACS_MEDIAN_HH_INC | Median Income | ACS |
| ACS_PER_CAPITA_INC | Per Capita Income | ACS |
| ACS_PCT_HS_GRADUATE | Percent High School Graduate | ACS |
| ACS_PCT_LT_HS | Percent Less Than High School Education | ACS |
| ACS_PCT_POSTHS_ED | Percent Post High School Education | ACS |
| ACS_PCT_HU_NO_VEH | No Vehicle Households | ACS |
| CEN_POPDENSITY_TRACT | Population Density | ACS |
| PCH_LACCESS_POP_10_15 | Low Access to Stores Difference Between 2010 and 2015 | Food Environmental Atlas |
| PCT_LACCESS_POP15 | Low Access to Stores | Food Environmental Atlas |
| Pct_alcohol | Percent Within Quarter Mile of | Modified Retail Food Environment |
|  | Alcohol Outlet | Index |
| Pct_store | Percent Within Quarter Mile of Healthy Food Store | Modified Retail Food Environment Index |
| Pct_park | Percent Within Half Mile of a Park | CDC |
| Pct_school | Percent Ages 5-9 Within Half Mile of Public Elementary School | CDC |
| Pct_no_pa | Percent Adults with No Physical Activity | CDC |
| Pct_smoking | Percent Adults that Smoke | CDC |
| Pct_bad_air | Concentration of 1-3-butadiene in the Air | CDC |
| EP_POV | Percent Below Poverty Estimate | Social Vulnerability Index |
| EP_UNEMP | Percent Unemployed | Social Vulnerability Index |
| EP_PCI | Per Capita Income | Social Vulnerability Index |
| EP_NOHSDP | Percent No High School Diploma | Social Vulnerability Index |
| EP_SNGPNT | Percent of Single Parent Households | Social Vulnerability Index |
| EP_MINRTY | Percent Minority | Social Vulnerability Index |
| EP_LIMENG | Percent with Poor English Fluency | Social Vulnerability Index |
| EP_MUNIT | Percent of Housing with at Least 10 Units | Social Vulnerability Index |
| EP_MOBILE | Percent of Mobile Homes | Social Vulnerability Index |
| EP_CROWD | Percent of Housing with More People than Rooms | Social Vulnerability Index |
| EP_NOVEH | Percent of Households with no Vehicle | Social Vulnerability Index |
| EP_GROUPQ | Percent in Group Quarters | Social Vulnerability Index |
| EP_UNINSUR | Percent Uninsured | Social Vulnerability Index |

### Data Analysis

My preliminary analyses included all of the census tracts in California and Illinois, but almost all of the variation in the data occurs in the metropolitan areas. Since health is largely local and all the interesting variation is in the cities, I pared the data down to the Chicagoland and Bay Area regions.

A Random Forest supervised machine learning model is used to identify the most important predictors of health in the different regions. Random forest is a predictive algorithm that creates a forest of random decision trees, each of which predicts a particular outcome. These trees are created using a variation of bagging on a subset of the entire set of independent variables. Randomness is incorporated to choose the subset of variables that identify the forking in each tree. Since only a subset is used in each of the trees of the random forest, the entire set of trees should be uncorrelated. The algorithm then takes the majority outcome of the trees, and uses the majority outcome as its prediction. Because the trees are created to have a low correlation, taking the majority outcome will result in a more accurate prediction than any individual tree could.

Instead of running random forest models on the entirety of California and Illinois, the analysis is limited to Chicago, and the Bay Area because there is a lot more variation within large cities, which is not reflected when analyzing the state as a whole. For each city, the analysis is comprised of a larger model which included all the factors included in the data set, and a smaller model, which included only the factors that can be more easily changed. These specific factors are especially interesting because they lend themselves more easily to understanding the impact of particular policies.

### Chicagoland

Chicagoland is a metropolitan region in the northeast corner of Illinois. There are regions of Chicagoland in Indiana and Wisconsin, but I analyzed specifically the Chicagoland area in Illinois. Chicagoland has approximately 9.5 million people, approximately 7.7 million of which live in Illinois. 6 counties make up the Chicagoland area in Illinois. These are Cook, Lake, Kane, Will, McHenry, and DuPage counties. These counties encompass 5,284 square miles, and include approximately 60.7% of Illinois’ population. There are 1,888 census tracts in the Chicagoland area.

In Chicagoland, the dependent variable, health level, is in the range from -2 to 2 based on the health value of that tract. My cutoffs for the 5 health levels were 132, 143, 159, and 175. So, if the health value of the tract is below 132, the health level is coded as 2. The higher the health value, the worse the health in the tract. Figure 3 shows a graph of the health values for the tracts in the Chicagoland area.

**Figure 3.**
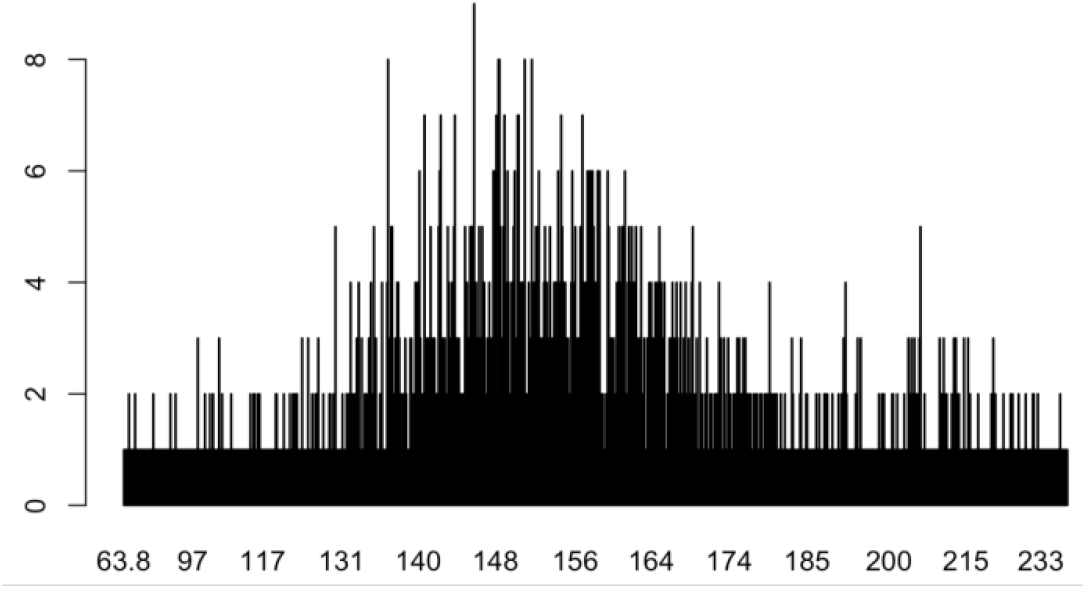
Chicagoland Health Distribution

The best predictors of health level are shown in Figure 4. This model explains 79.02% of the variance in the data. Here, the longer the line, the greater the percentage decrease in mean squared error for the prediction. As we can see from the figure, the variables that are the best predictors of health are population density and Black percentage of the population. This tells us that neighborhoods that are densely populated tend to be less healthy, and that areas with higher concentrations of Black residents have the worse health outcomes in the Chicagoland area. This pattern is not particularly surprising since White and Black racial issues have a long history in Chicago. Black inequality has been a long-embedded issue in Chicago, which has been caused by a history of structural racism. In the 1900s, the Black population in Chicago faced heavy restrictions on where they could live and how they could buy homes. They were not offered traditional mortgages, and the discrimination caused the Blacks that could afford to, to leave. As a result, this caused the Black neighborhoods that remained to be poorer and have significantly less resources. Having less resources meant they lacked schools, grocery stores, health care facilities, and healthy air, which resulted in poorer health for those that lived there (Gross, 2021). Unfortunately, the effects of this segregation are very hard to change, and its effects are still seen in Chicago today. From 1990 to 2012 the average household income for White households has increased by over 22000 USD, but in this same time frame, the average household income has actually *decreased* for Black households (Grabinsky and Reeves, 2015).

**Figure 4.**
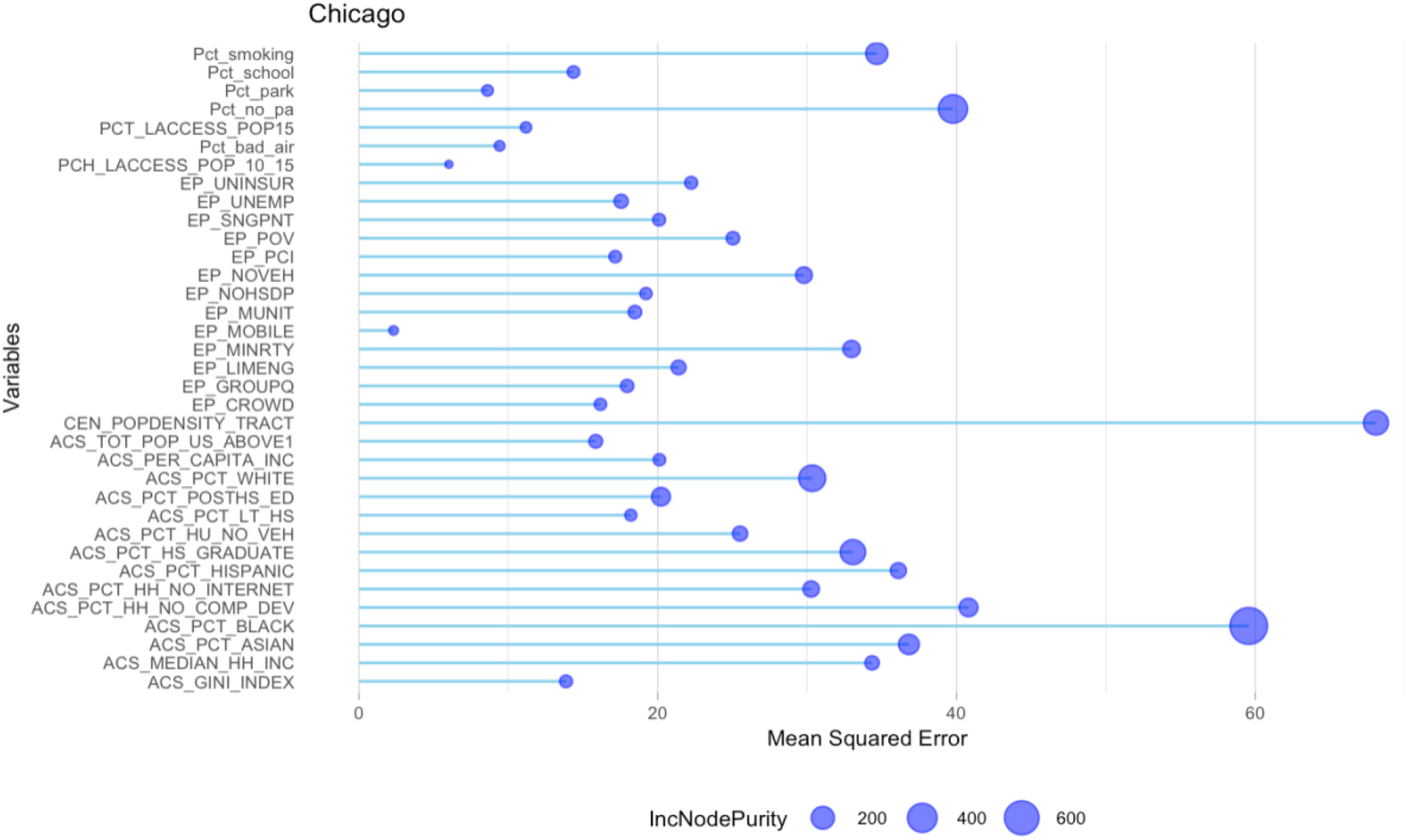
Predictors of Health in Chicagoland.

Many factors, though not all of the factors that are related to poor health areas, are difficult to change. There are some factors, however, that are more easily changed with policy changes. For example, it would be easier to alter proximity to schools, the location of parks and stores, access to a computing device and the internet, or the quality of the air. While these factors are not easy to change either, there is a clearer path for how one might change them. For the factors that are more easily changed, I conducted a separate random forest analysis. Figure 5 shows the best predictors from a random forest model but now the random forest model includes only variables that perhaps can be changed more easily by enacting various policies. This model explains 38.34% of the variance in the data, which is nearly half of the variance explained by the larger model.

**Figure 5.**
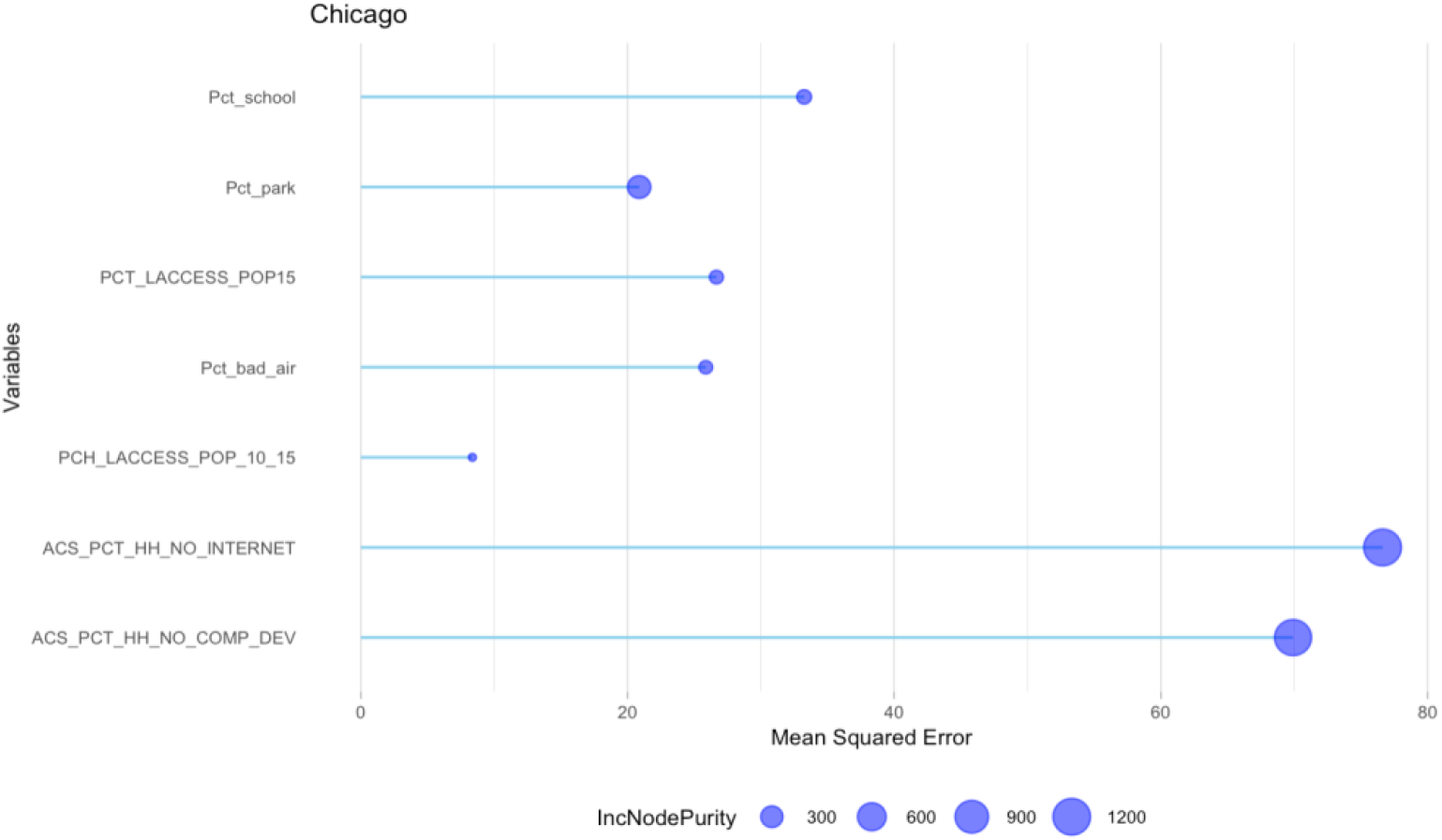
More Easily Changeable Predictors of Health in Chicagoland

Out of these factors that we could change, the percentage of households without internet access, and the percentage of households without access to a computing device are by far the bigger predictors of healthy neighborhoods. These two factors are similar, and together measure access to information. Having a computer and internet access is likely so important for health because we gain so much information from the internet. Households without internet access are likely less knowledgeable about how to keep themselves healthy. In addition, the internet is useful when you are potentially unwell, as it allows you to look up your symptoms to see potential causes, as well as how best to recover.

### Bay Area

The Bay Area is an extensive metropolitan region in northern California. There are approximately 7.76 million people living in the Bay Area, which make up approximately 20% of California’s population. There are 9 countries that make up the Bay Area. These are Alameda, Contra Costa, Marin, Napa, San Francisco, San Mateo, Santa Clara, Solano, and Sonoma counties. These counties encompass 6,966 square miles, which cover 1,402 census tracts.

For the Bay Area, the dependent variable, health level, is in the range from -2 to 2 based on the health value of that tract. My cutoffs for the 5 health levels were 123, 132, 144, and 153. So, if the health value of the tract is below 123, the health level is coded as 2. The higher the health value, the worse the health in the tract. Figure 6 shows a graph of the health values for the tracts in the Bay Area.

**Figure 6.**
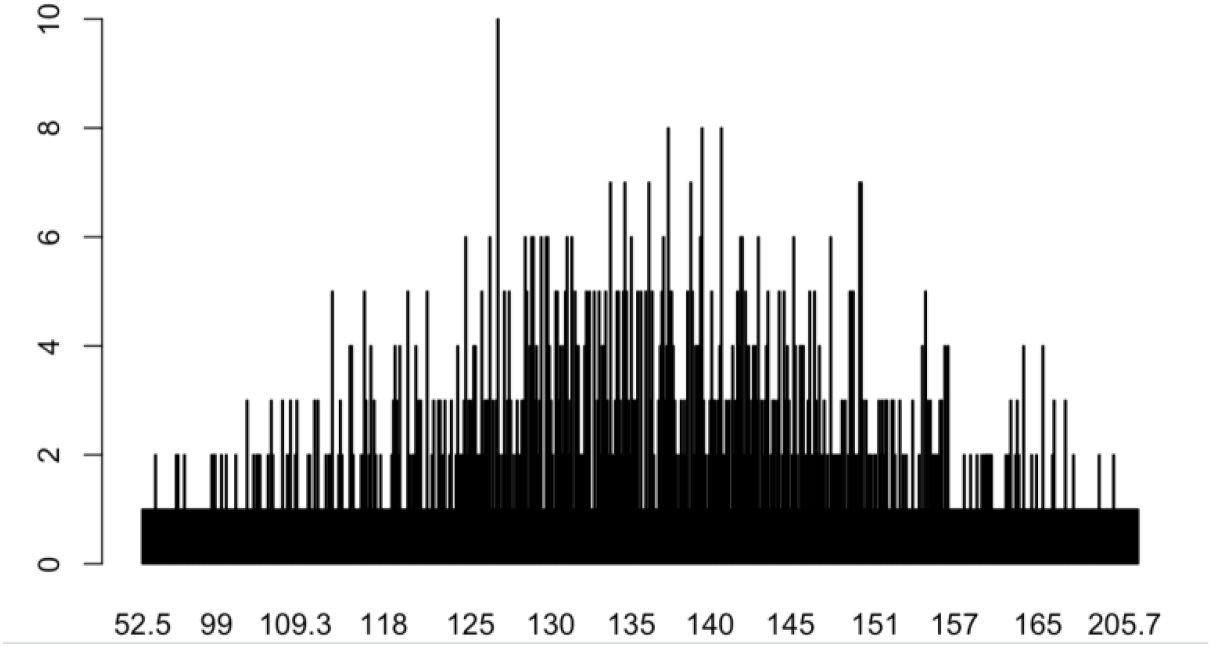
Bay Area Health Distribution

The best predictors of health level are shown in Figure 7. This model explains 66.51% of the variance in the data. As we can see from the figure, there are many important factors, especially race, as the percentage of White, Black, Hispanic, and Asian people are all important predictors. A regression model shows that higher concentrations of Blacks correlate to bad health, and higher concentrations of Whites and Asians correlate to better health. This explanation is too simplistic, however, and is misleading. This is because the Bay Area has some of the highest levels of income inequality in the country. In 2018, the 90th percentile of Bay Area residents earned more than 12 times as much as those in the bottom 10th percentile (Hellerstein, 2020).

**Figure 7.**
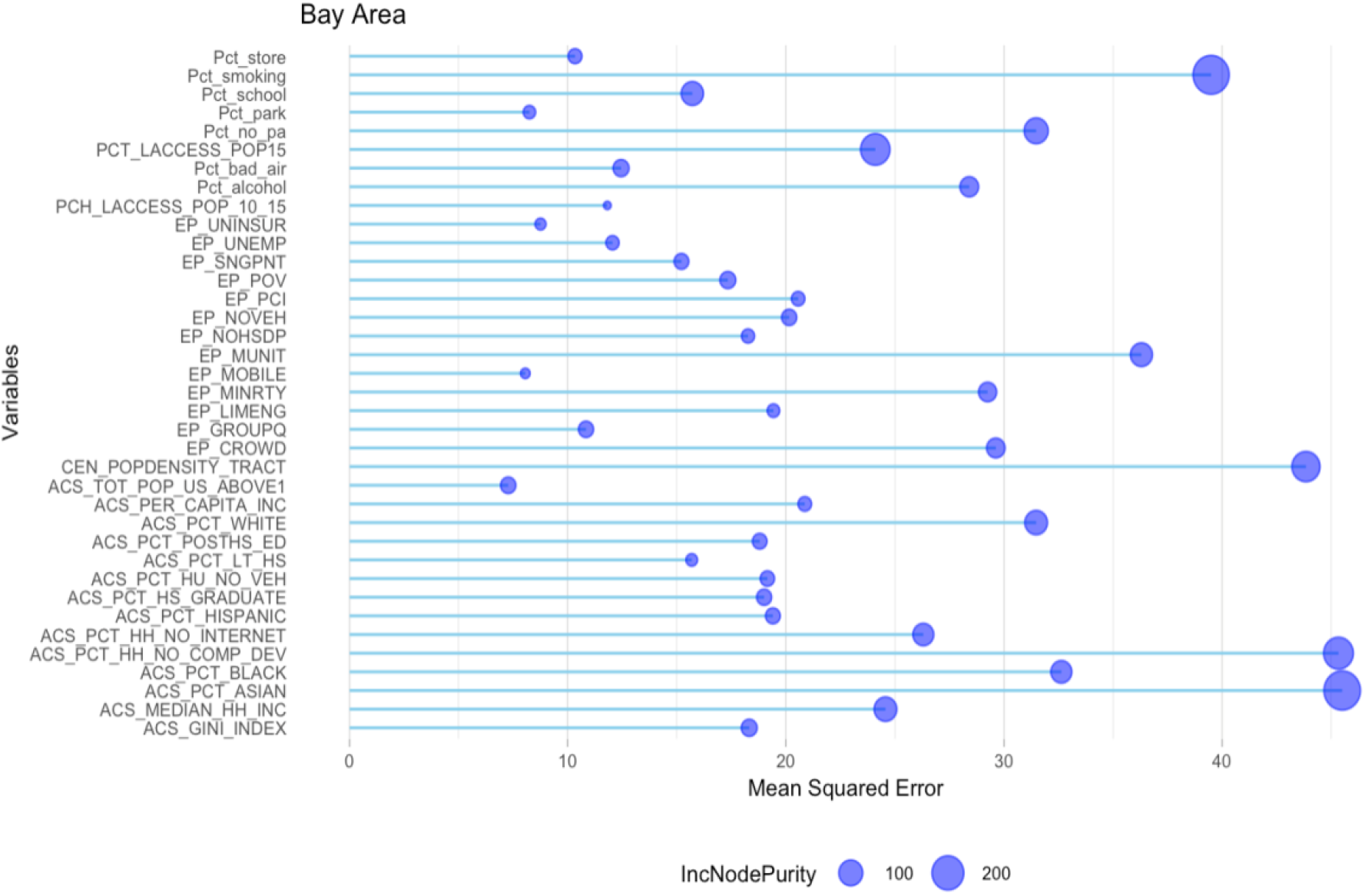
Predictors of Health in the Bay Area

Inequality is an issue which transcends race, as there are very healthy, and very unhealthy people in each race. This can be seen in the plots comparing each race against the health variable. In Figure 8, which graphs health against percent White, we can see that the unhealthiest tracts are ones which have a very small percentage of Whites *and* the ones which have a very large percentage of Whites. This means that having a higher percentage of Whites does not imply that a tract will be healthier, and in fact some of the highest percentages also have the worst health.

**Figure 8.**
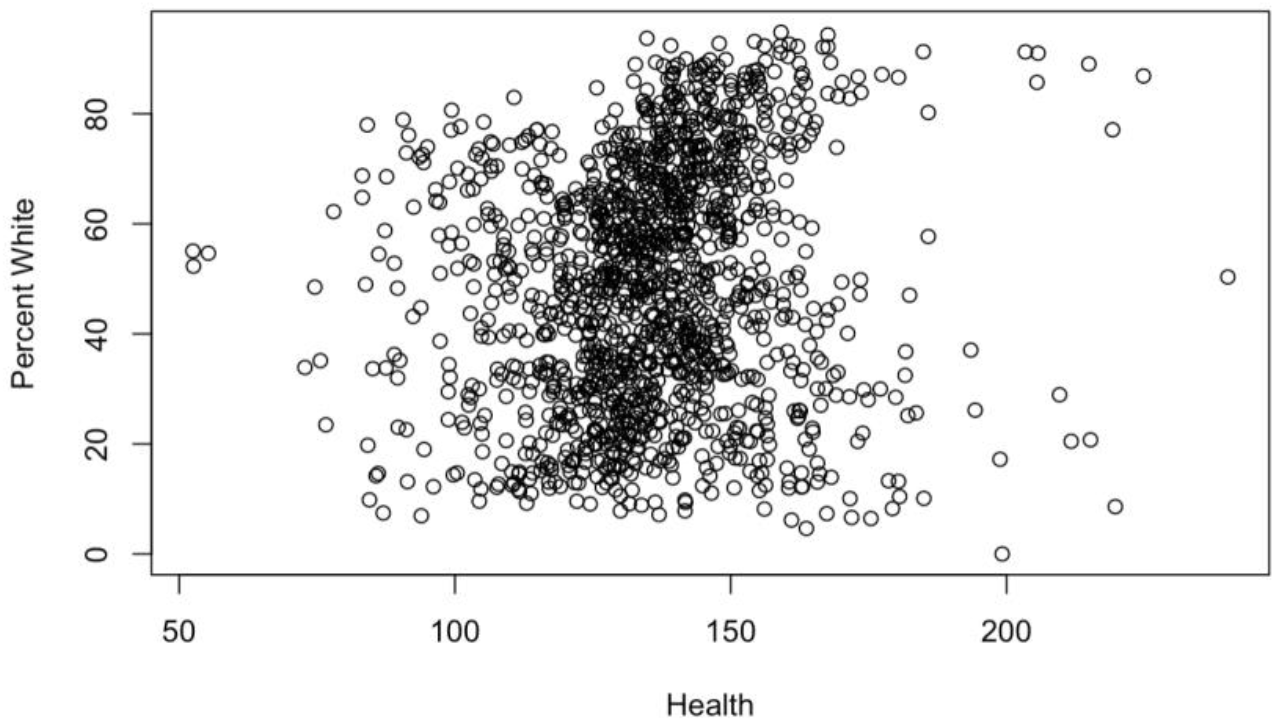
Health and Percent White

A similar result is also found in the graphs comparing Asians and Whites with health. Figure 9 plots health against percent Asian. In this plot, we can similarly see that despite higher percentage of Asians corresponding to better health *in general*, the tracts with the worst health both have low percentages of Asians, and high percentages of Asians. This is because there is a large amount of inequality among Asians in the Bay Area. There are some Asians that are very rich, but there are also Asians that are very poor. This is illustrated with the two plots in Figure 10. The map on the left shows poverty in the city of San Francisco. The map on the right shows percent Asian in the city of San Francisco. We can see that the places with the highest densities of Asians are either very wealthy, or very poor. A similar result, though not quite as striking as for Asians, is seen among Blacks as well, as shown in Figure 11. Overall, in the Bay Area, we can see that there is an underlying issue of inequality that transcends race.

**Figure 9.**
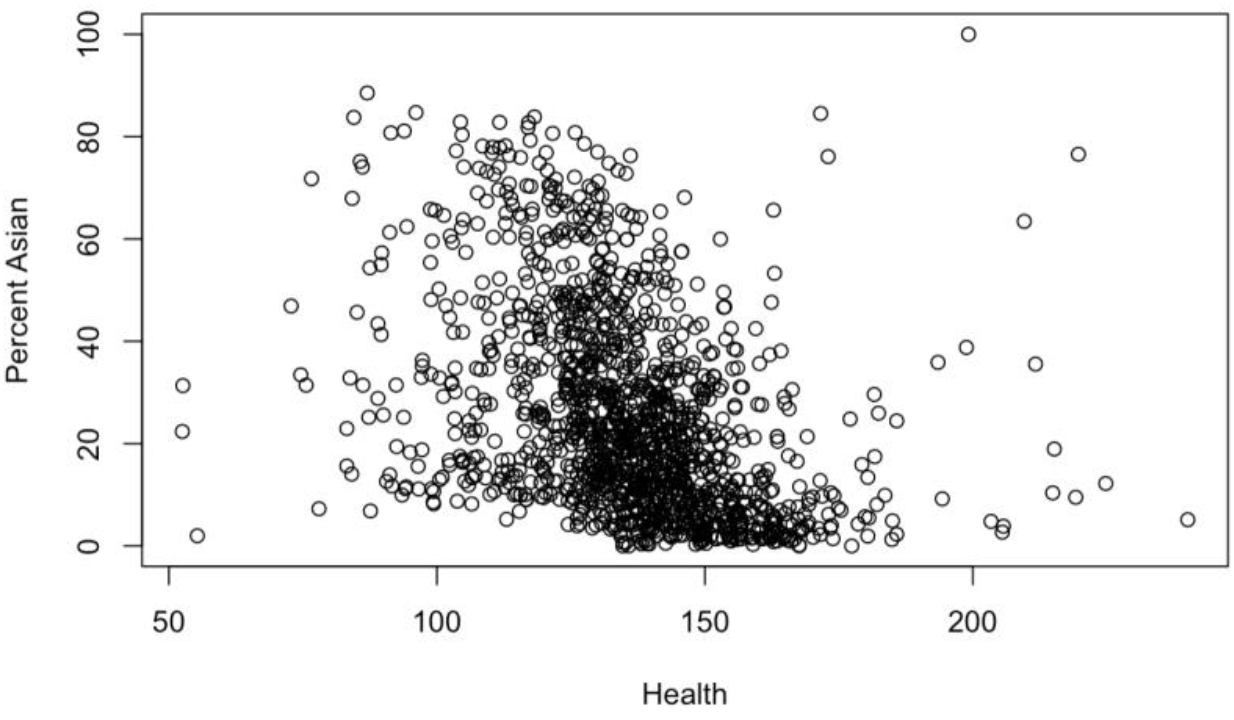
Health and Percent Asian

**Figure 10.**
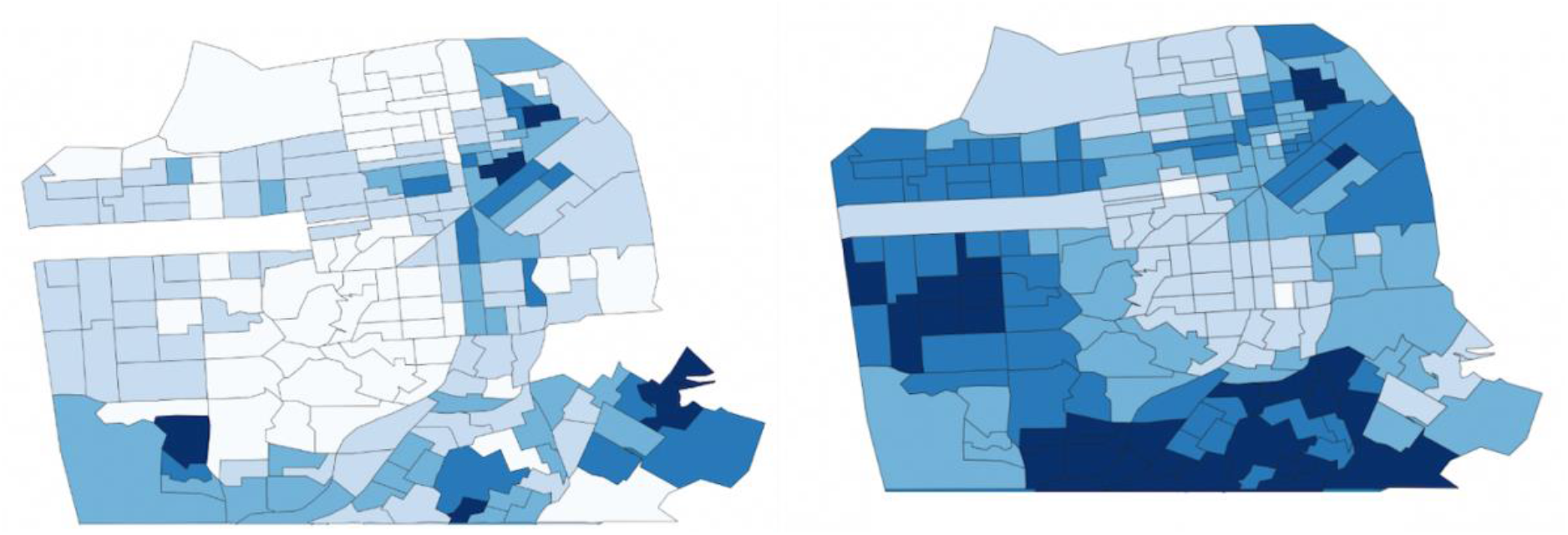
Percent Poverty (left) and Percent Asian (right) in San Francisco

**Figure 11.**
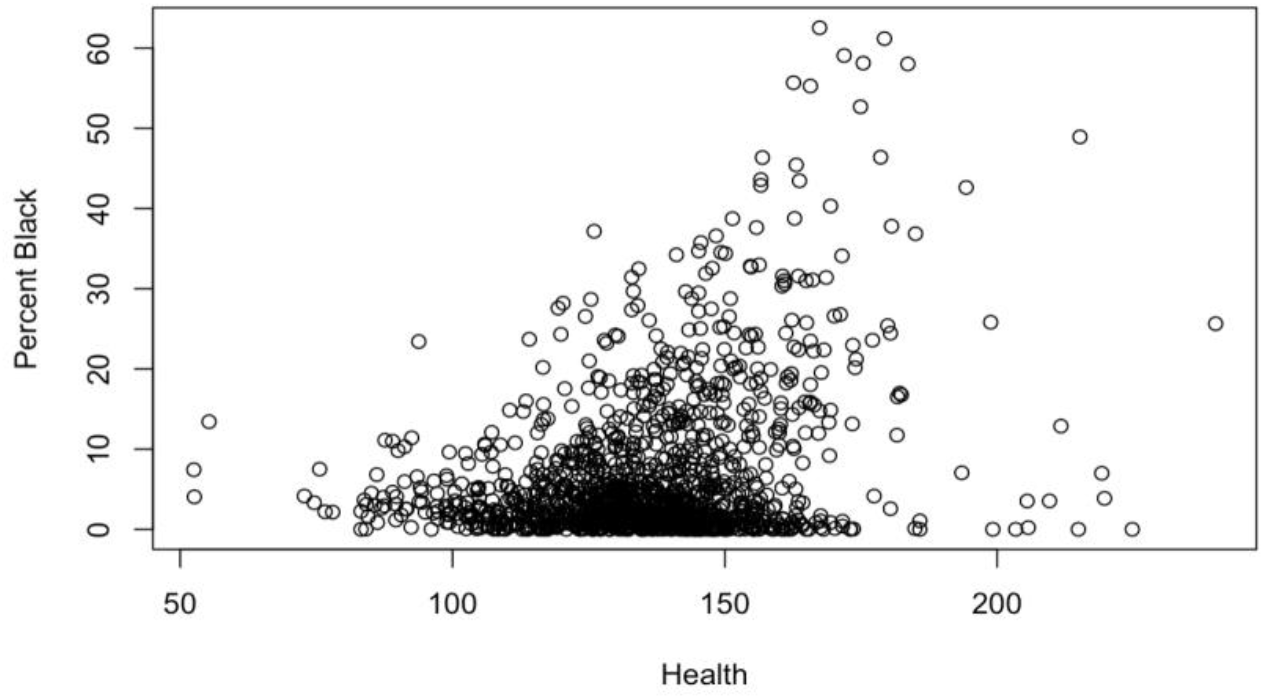
Health and Percent Black

For the factors that are more easily changed, a separate random forest analysis is conducted for the Bay Area. This model explains 48.21% of the variance in the data, which is more than two thirds of the variance explained by the larger model. Figure 12 includes the same factors from Figure 5 with the inclusion of population percentage within a quarter mile of an alcohol outlet, and population percentage within a quarter mile of a healthy food store. These factors for the Bay Area analysis are included because there was more available data for California than was available for Illinois. Out of these factors, no computing device, no internet, proximity to alcohol outlets, proximity to healthy food stores, and proximity to schools are the most predictive.

**Figure 12.**
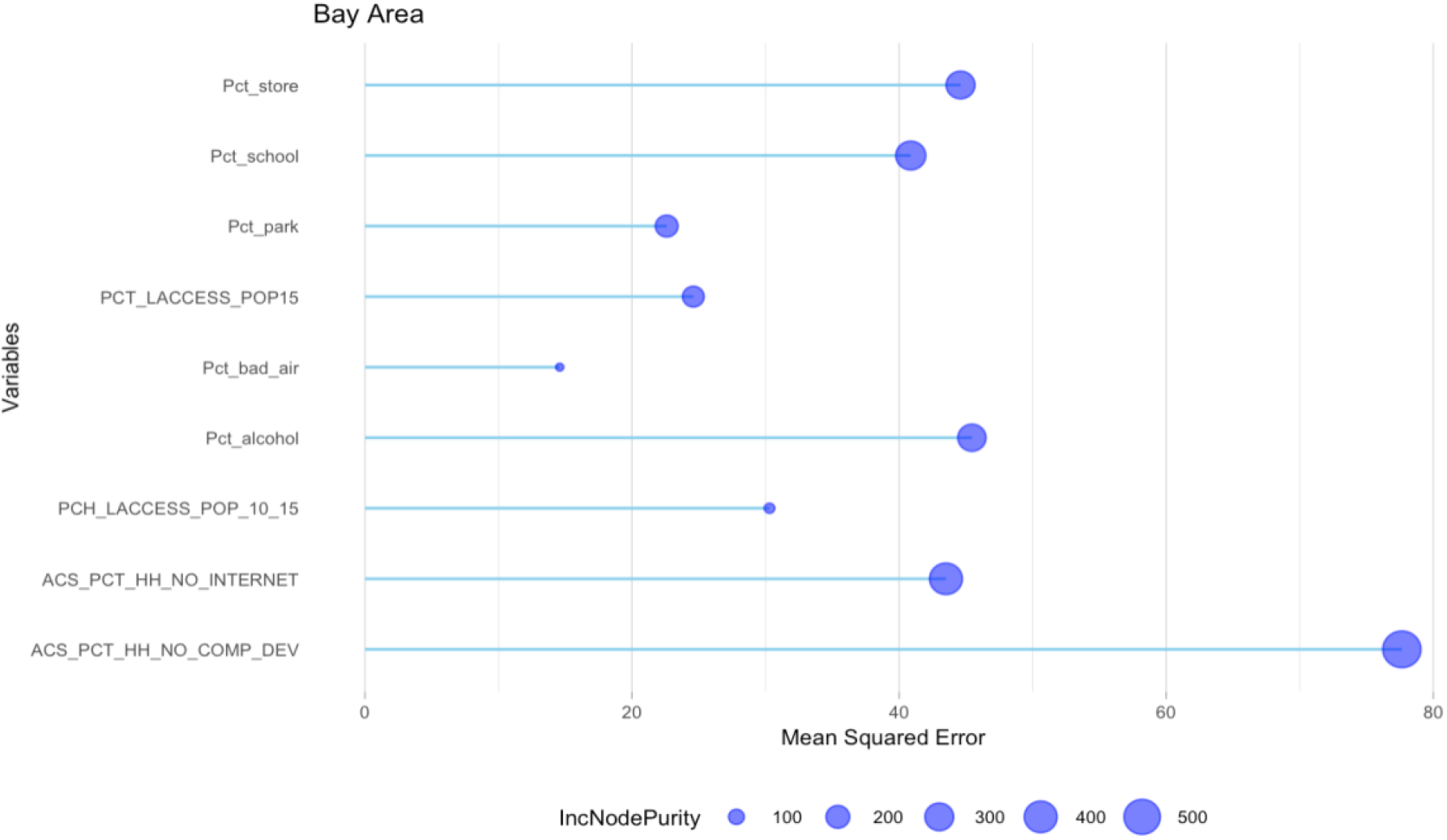
More Easily Changeable Predictors of Health in the Bay Area

No computing device and no access to internet are important in both the Bay Area and Chicagoland models, and are likely important for a similar reason. Having access to information is an important factor to maintaining good health. The percentage of kids within close proximity to a school is also an important predictor. This could be related to health for many different reasons. For example, if a kid needs to walk farther to school, they would need to wake up earlier, which impacts their sleep, which in turn impacts their health. Additionally, walking farther to school could mean worse exposure to bad environment factors, such as pollution. If a kid needs to walk farther to school, that could mean they breathe in more polluted air every day, which would impact their health. Having close proximity to healthy food stores, and not having close proximity to alcohol outlets are also important. This is likely because we frequently go to places that are close to us, and being able to access healthy food helps maintain health, while drinking alcohol does the opposite. If there are no healthy food stores close by, it is much more difficult to eat well, and people would instead be forced to settle for less healthy options. Similarly for alcohol, having easily accessible alcohol outlets is bad for a neighborhood, because it will cause people to drink more frequently. This means that it would be helpful to remove drinking outlets from unhealthy neighborhoods, as well as making sure everyone has a way to access healthy food.

## Discussion and Conclusion

In the U.S. there are many health disparities that stem from structural historical factors that are very difficult to change. For example, some factors such as race, income, age, and gender, are not easily changed, even if we know they are significant predictors of health disparities. This can be seen with the Black population in Chicago. Racial health disparity for Blacks and general inequality have existed for a very long time, with some but little progress being made in the last hundred years. Although we do need to work to improve these issues, we know from the long history that this change will not be easy. As such, I focused on things that we can work on changing that would still be effective, yet be easier and more straightforward to implement.

One important predictor of areas with poor health in both Chicago and the Bay Area is lack of internet access and lack of access to a computing device. This means that providing internet access to as many households as possible will improve health outcomes. One way this might be accomplished is by installing municipal wireless networks in unhealthy areas. This would allow those people living in those areas to have free or cheap access to the internet. It is also important to provide people with access to computing devices. One way we could work toward this goal is by having public schools give their students cheap computers to use at home. Alternatively, improvement could be approached indirectly, by installing computers for public use in unhealthy areas. Computers could be put in public libraries, or public schools, which might lessen the negative impact for households that do not have a computing device.

In the Bay Area, proximity to schools, healthy food stores, and alcohol outlets, are all predictors of healthy areas. One way we could attempt to reduce alcohol outlets proximity is by creating restrictions on alcohol outlet locations, or on the number of alcohol outlets in an area. This would need to be done by state laws. If we had such laws, this could decrease how readily accessible alcohol outlets are, which could reduce the amount of drinking in that area. To increase proximity to schools and healthy food stores within a region, we could work towards constructing more schools and stores in locations in more geographically equitable ways. This would be helpful in regions with either a lack of schools/stores, or poorly located schools/stores. Some of these issues can be addressed by city planners or through city ordinances that restrict what can be built where in the city.

Overall, health has become an increasingly important subject since the pandemic. The pandemic has increased our concern for health, and has changed our perspective on what it means to be healthy. Previously, we might have viewed health disparities as an unfortunate issue for those who are less privileged, but the pandemic has taught us that we are all interconnected. Health is not an individual factor, but a factor that depends heavily on the community around us. Reducing health disparities should be a priority for everyone, as it improves health not just for those who are less fortunate, but improves health for society as a whole.

## Data Availability

All data produced in the present work are contained in the manuscript

## Footnotes

1 The VA (value-added) metric is a measure of the quality of a teacher, calculated based on a teacher’s impact on student test scores.

